# Short-term antibody response and tolerability after one dose of BNT162b2 vaccine in patients receiving hemodialysis: A report from the Quebec Renal Network COVID-19 study

**DOI:** 10.1101/2021.03.30.21254652

**Authors:** Rémi Goupil, Mehdi Benlarbi, William Beaubien-Souligny, Annie-Claire Nadeau-Fredette, Debashree Chatterjee, Guillaume Goyette, Caroline Lamarche, Alexander Tom, Andrés Finzi, Rita S. Suri, on behalf of the Réseau Rénal Québécois (Quebec Renal Network) COVID-19 Study Investigators

**Author notes:** **Correspondence:** Rita S. Suri, Director, Division of Nephrology, Associate Professor of Medicine, McGill University, Room D05.7166,McGill University Health Centre, 1001 Decarie Blvd, Montreal, QC H4A 3J1, T., F. co-first author. co-senior author. **Disclosures :** None.

## Abstract

**Background:** Patients receiving in-center hemodialysis (HD) are at high risk of exposure to SARS-CoV-2 with high mortality, and response to vaccination is uncertain.

**Methods:** We obtained serial plasma from 58 HD patients and 32 health-care workers (HCW) before and after vaccination with one dose of the BNT162b2 mRNA vaccine; as well as convalescent plasma from 11 HD patients who survived COVID-19. Anti-RBD (region binding domain of the SARS-CoV-2 Spike protein) IgG and IgM levels were measured by ELISA. Groups were stratified by evidence of prior SARS-CoV-2 infection.

**Results:** In HD patients without prior SARS-CoV-2, antiRBD levels were significantly lower at 4 and 8 weeks after vaccination, compared to in HCWs after 3 weeks (p<0.001), and to convalescent plasma (p=0.002). Anti-RBD IgG was non-detectable in 29/46 (63%) of HD, compared to 1/16 (6%) of HCWs (p<0.0001). No patient with non-detectable levels at 4 weeks developed antiRBD by 8 weeks. In HD patients with prior SARS-CoV-2, mean 8-week anti-RBD IgG levels were similar to controls at 3 weeks (p=0.16), and to convalescent plasma (p=0.45). No patients reported symptoms 7 days after vaccination on a standardized questionnaire.

**Interpretation:** While the BNT162b2 vaccine was well-tolerated in hemodialysis patients, a single dose failed to elicit a humoral immune response in the majority of SARS-CoV-2 naïve patients even after prolonged observation. In those with prior SARS-CoV-2 infection, the humoral response after vaccination was delayed. Whether HD patients develop T-cell responses requires further study. Until then, we advise the second dose be administered to all HD patients at the recommended 3-week time interval, and that rigorous SARS-CoV-2 infection prevention and control measures be continued in dialysis units until vaccine efficacy is proven.

## Introduction

Patients with end-stage kidney disease receiving in-center hemodialysis have been uniquely vulnerable during the COVID-19 pandemic. Unlike for other people, self-isolation to avoid exposure to SARS-CoV-2 is impossible. Most HD patients must leave their homes 3 times weekly to receive their life-saving treatments, often in shared spaces for hours at a time. Once infected, case fatality rates are 20-30% - 10 times higher than in the general population(1, 2). Advanced age, multiple comorbidities and blunted immune response likely all contribute to the high COVID-19 death rates in hemodialysis. Some centers have thus prioritized hemodialysis patients for vaccination.

To facilitate wider vaccine distribution during current shortages(3), the National Advisory Committee on Immunization (NACI) of Canada has recommended delaying the second dose of the BNT162b2 vaccine from 3 to 16 weeks(4). The reported clinical efficacy of the BNT162b2 was reported to be >80% at 3 weeks the first dose in a randomized trial(5). However, hemodialysis patients were not enrolled in this trial(5). Many HD patients have impairments in both humoral and cellular immune responses(6), and are noted to have lower antibody responses to other vaccines(7). Thus, whether HD patients develop a robust immune response after vaccination against SARS-CoV-2 is uncertain.

In this context, we sought to determine if short-term antibody response is comparable between hemodialysis patients and healthcare workers (HCWs) vaccinated with a single dose of the BNT162b2 mRNA vaccine, and how this compares to antibody responses in dialysis patients who survived natural infection with SARS-CoV-2.

## Methods

We have been following in-center hemodialysis patients in the Réseau Rénal Québecois COVID-19 study since March 2020, with participation of 54 of Quebec’s 55 hemodialysis units. Patients dialyzing in 5 academic centers were recruited for this vaccine substudy. Here we report 8-week data on the first 58 patients who were vaccinated the week of Jan 26^th^ 2021. This study was approved by the local research ethics board and all patients provided written informed consent.

### Protocol

We obtained clinical data and serial plasma samples from 58 hemodialysis patients before and every 4 weeks after vaccination with the BNT162b2 (Pfizer-BioNTech) vaccine. All patients completed a symptoms questionnaire 7 days after vaccination. These patients have been prospectively followed for symptoms of COVID-19 using a systematic screening questionnaire at each hemodialysis treatment session since March 200. We also performed weekly surveillance SARS-CoV-2 testing by PCR of nasopharyngeal swabs from April 2020 to July 2020, and from October 2020 until now.

### Control Patients

Health-care workers who received the BNT162b2 vaccine served as controls with plasma obtained before, and 3 weeks post vaccination (12 week samples are pending in this group). In addition, we obtained convalescent plasma from hemodialysis patient survivors 4-12 weeks after PCR-confirmed SARS-CoV-2 infection.

### Antibody Measures

IgG antibodies against the receptor binding domain (RBD) of SARS-CoV-2 Spike glycoprotein were assessed in plasma samples using an enzyme-linked immunosorbent assay and IgG levels reported as relative light units (RLU) normalized to CR3022 mAb, as described(8, 9) IgG and IgM antibodies against the receptor binding domain (RBD) of SARS-CoV-2 Spike glycoprotein were assessed in plasma samples using an enzyme-linked immunosorbent assay (ELISA)(8, 9). Bovine serum albumin and CR3022 monoclonal antibody (mAb) against SARS-CoV-2 S glycoprotein were used as negative and positive controls, respectively. Anti-RBD IgG and IgM levels were reported as relative light units (RLU) normalized to CR3022 mAb. Seropositive threshold was established as mean RLU from COVID-19 negative plasma obtained from 10 volunteers pre-pandemic plus 3 standard deviations of this mean(8, 9). We have previously shown excellent reliability of this assay in detecting anti-RBD IgG levels in a cross-sectional cohort of infected individuals: anti-RBD IgG levels were detectable with this assay in 85% of infected individuals at 14 days, and in 100% by 53 days after symptom onset(8). Finally, we measured anti-nucleocapsid IgG (anti-N, ELISA, Abbott Architect 1200SR) to rule out new asymptomatic natural infection after vaccination.

### Statistical Analysis

We stratified groups by past COVID-19 infection (positive PCR to SARS-CoV-2 *or* positive anti-RBD before vaccination). For one dialysis patient in whom an 8 week sample could not be obtained, the 4-week value was carried forward. Mean RLUs were compared using student’s t-test, and the proportion of patients with non-detectable anti-RBD IgG levels were compared with chi-square test. Two-sided p-values of <0.01 were considered significant, considering multiple comparisons.

## Results

Dialysis patients were considerably older than HCWs, with a high rate of diabetes (table 1). No dialysis patient reported symptoms within 7 days after vaccination, and none developed SARS-CoV-2 infection after vaccination as defined by presence of new anti-N antibody or serial PCR. Anti-RBD levels are provided in figure 1 legend.

**Table 1:** Baseline Characteristics.

| Variable | Vaccinated, COVID-19 naïve |  | Vaccinated, Prior COVID-19 |  | Unvaccinated<sup>a</sup> |
| --- | --- | --- | --- | --- | --- |
|  | HCWs<br>[Group H1,<br>N=16] | DIALYSIS<br>[Group D1,<br>N=46] | HCWs<br>[Group H2,<br>N=16] | DIALYSIS<br>[Group D2,<br>N=12] | DIALYSIS<br>[Group D3,<br>N=11] |
| Age (yrs) | 48 (10.1)<br>[52, 21-59] | 74.4 (12.4)<br>[77, 41-92] | 47 (12.8)<br>[46, 23-65] | 80.3 (9.7)<br>[84, 56-90] | 65.4 (17.1)<br>[70.5, 35-88] |
| Female sex (N, %) | 81 | 21 (46) | 50 | 7 (58) | 6 (55) |
| Long-term Care (N, %) | 0 | 2 (4) | 0 | 2 (17) | 0 |
| Immunosuppressive medications (N, %) | 0 | 5 (11) | 0 | 1 (8) | 2 (18) |
| Past Kidney Transplant (N, %) | 0 | 0 | 0 | 0 | 3 (27) |
| Diabetes (N, %) | 0 | 28 (61) | 0 | 6 (50) | 7 (63) |
| Days since COVID-19 infection<sup>b, c</sup> | n/a | n/a | 249 (58.97)<br>[266, 116-309] | 276 (30)<br>[285, 192-292] | 82 (22)<br>[88, 55-121] |
| Days since vaccination | 22 (3.29)<br>[21, 16-26] | 28 (1.5)<br>[28, 26-31] | 20 (1.85)<br>[21, 17-25] | 27.5 (2.0)<br>[27, 26-31] | n/a |
| Hospitalized for COVID-19 | n/a | n/a | n/a | n/a | 6 (55) |
| Intensive care | n/a | n/a | n/a | n/a | 1 (9) |

<sup>a</sup>Defined as PCR positive test in dialysis cohort, and onset of symptoms in HCW cohort
<sup>b</sup>For 2/12 dialysis patients, time of infection was unknown as they were diagnosed by serology only.
<sup>c</sup>At the time of writing, all patients in this group had survived (min 55 days)
Data are presented as mean (SD), [median, range] unless otherwise specified.

**Figure 1:**
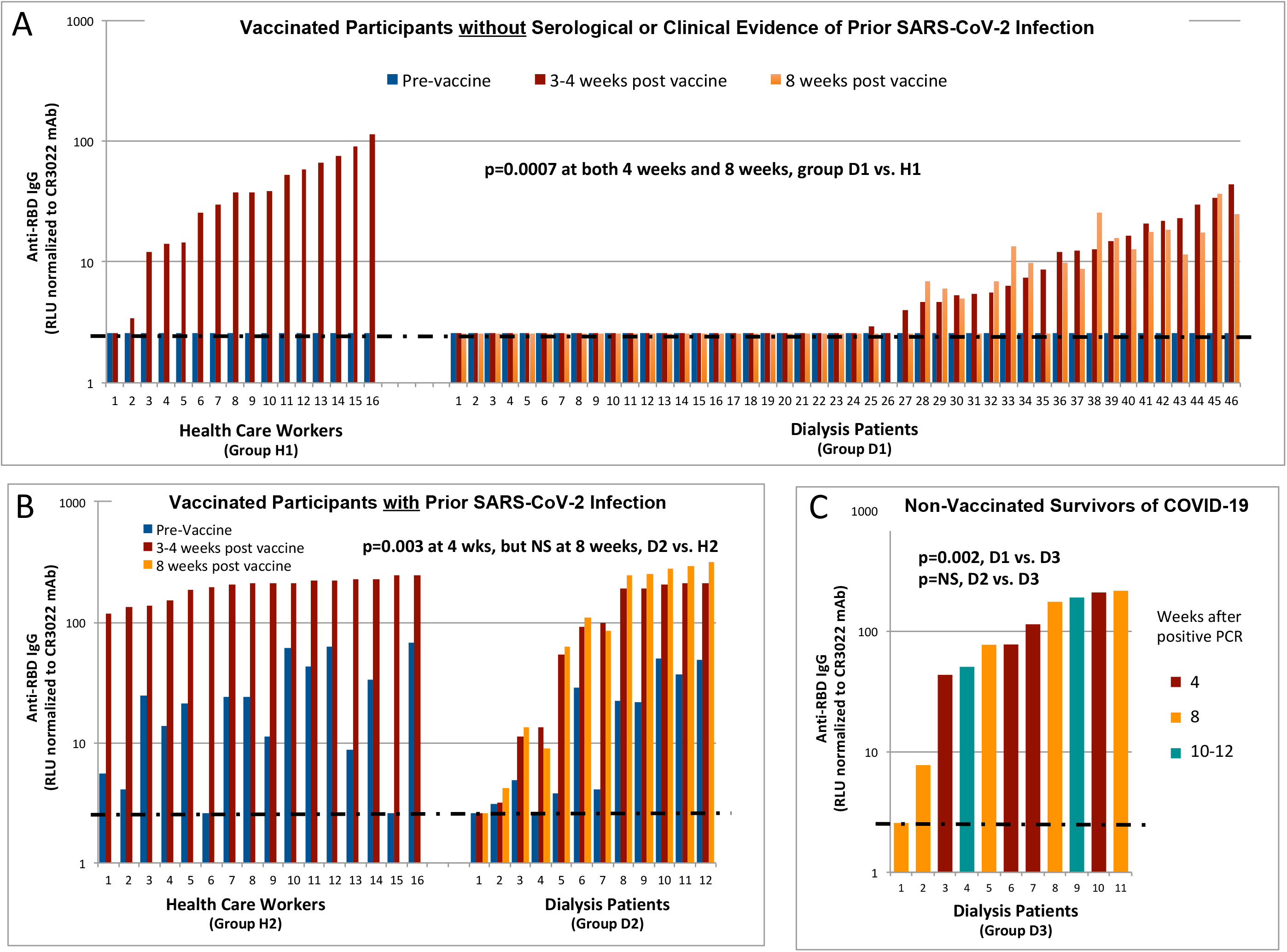
Anti-RBD IgG in Dialysis Patients and Health Care Workers. Each bar represents a different individual. Post-vaccine samples were taken 21 days (range 16-26) after a single dose of BNT162b2 mRNA vaccination in health-care workers, and after 28 days (range 26-31) and 56 days (range 54-59) in dialysis patients. Anti-RBD IgG levels were measured using ELISA, subtracting the signal obtained with bovine serum albumin, and normalized to the signal obtained with CR3022 monoclonal antibody to SARS-CoV-2 in each plate. Dotted line represents the “positive threshold for detection” of anti-RBD IgG, defined as the mean RLU plus 3 standard deviations from plasma samples obtained from ten volunteers before the pandemic. RLU = relative light units. **A:** Anti-RBD IgG levels in COVID-19 naïve vaccinated dialysis patients (n=46) and COVID-19 naïve vaccinated health care workers (n=16). Group H1: 42 ± 32 RLU at 3 weeks; Group D1: 7.9 ± 9.5 RLU at 4 weeks (p=0.0007 vs. H1); Group D1: 7.1 ± 7.8 RLU at 8 weeks (p = 0.0006 vs. H1). No antibody response was observed in 1/16 (6%) of health-care workers at 3 weeks, and 29/46 (54%) of dialysis patients at 8 weeks (p<0.0001). **B:** Anti-RBD IgG levels in vaccinated dialysis patients (n=12) and vaccinated health care workers (n=16), each with evidence of prior COVID-19 infection Group H2: 196 ± 41 RLU at 3 weeks; Group D2: 108 ± 90 at 4 weeks (p=0.0031 vs. H2): 139 ± 127 RLU at 8 weeks (p=0.155 vs. H2). **C:** Anti-RBD IgG levels in convalescent plasma obtained from dialysis patients (n=11) between 4-12 weeks after COVID-19 infection. Group D3: 105 ± 79 [p = 0.002 vs. D1 at both 4 and 8 weeks; p= NS vs. D2 at both 4 and at 8 weeks].

### SARS-CoV-2 naïve group

Dialysis patients without prior COVID-19 exhibited significantly lower anti-RBD IgG levels at both 4 and 8 weeks after vaccination, compared to HCWs at 3 weeks (figure 1, panel A), and compared to convalescent plasma from dialysis patient survivors of COVID-19 (figure 1, panel C). The number of dialysis patients with non-detectable antiRBD was 26/46 at 4 weeks (56%, 95% CI = 42-71%), and 29/46 at 8 weeks (63%, 95% CI 49-77%), compared to 1/16 HCWs at 3 weeks (6%, 95% CI 0-18%); (p<0.001 for 4 week comparison, and <0.0001 for 8 week comparison). Importantly, none of the 26 dialysis patients with non-detectable antibodies at 4 weeks had developed detectable antiRBD by 8 weeks. In those with detectable levels at 4 weeks, mean anti-RBD IgG was still significantly lower in dialysis patients than in HCWs (14.7±11.1 vs. 44.8±31.8 RLU, p=0.0046), and this did not increase by 8 weeks.

### Prior SARS-CoV-2 group

Twelve dialysis patients had evidence of prior SARS-CoV-2 infection (8 with both positive PCR and anti-RBD, 2 with positive PCR only, and 2 with positive anti-RBD IgG). These 12 patients developed anti-RBD IgG levels 4 weeks after vaccination that were significantly lower than those of HCW controls (figure 1, panel B), but not significantly different than those in convalescent plasma (figure 1, panel C). By 8 weeks, anti-RBD IgG levels in these 12 patients had increased to levels comparable to those in HCWs at 3 weeks (p=0.16). The levels were below detectable levels in 1/12 dialysis patients compared to 0/16 HCWs (p=NS).

### Anti-RBD IgM

There was no difference in anti-RBD IgM levels at 4 weeks between vaccinated HCW and dialysis patients, either in individuals without prior SARS-CoV-2 (HCW 4.9 ± 4.6 RLU, dialysis 3.8±1.1 RLU, p=0.34), or with prior SARS-CoV-2 (HCW 10.5 ± 14.8 RLU, dialysis 12.1 ± 12.2 RLU, p=0.67).

## Interpretation

In a randomized trial, the BNT162b2 vaccine was demonstrated to be >80% clinically effective by 3 weeks after the 1^st^ dose in healthy individuals(5). Indeed, all but one of our HCWs developed anti-RBD IgG within 3 weeks of a single dose of this vaccine. In contrast, we found that COVID-19 naïve patients receiving hemodialysis showed lower anti-RBD IgG levels than healthy controls after a single dose of the BNT162b2 vaccine, with more than 50% of patients failing to seroconvert at 4 weeks. Importantly, dialysis patients who did not respond at 4 weeks remained non-responders at 8 weeks, arguing against the possibility of a delayed response in these individuals. Humoral response was better after single dose vaccination in dialysis patients who had been previously infected with COVID-19, with mean anti-RBD IgG levels similar to healthy controls by 8 weeks.

These results suggest that one dose of the BNT162b2 vaccine is insufficient to elicit a robust humoral immune response in the majority of dialysis patients who have never been previously exposed to SARS-CoV-2, casting doubt on whether a single dose is effective in this population. This is consistent with prior studies that show that dialysis patients may require higher or repeated doses of hepatitis B vaccine to elicit a response(7). We do not know if anti-RBD IgG confers immunity against SARS-CoV-2, but it is likely a good surrogate as we have previously shown that anti-RBD IgG response correlates with F_c_ mediated effector functions and cellular responses(10), and all survivors of COVID-19 infection in our study mounted an anti-RBD response 4 to 12 weeks after infection. Further, while we cannot rule out that vaccinated dialysis patients with undetectable anti-RBD IgG may have developed protective cellular immune responses, in previous studies, neutralization, F_c_ function, and SARS-CoV-2 specific T cell responses have only been observed in individuals who elicited RBD-specific antibodies(10, 11). Whether a second dose would induce a more robust humoral response in dialysis patients without previous SARS-CoV-2 is uncertain. In another study, 95% (54/56) patients developed detectable anti-Spike IgG levels 30 days after a second dose of the BNT162b2 vaccine, but we do not know how many of these patients were previously infected with SARS-CoV-2(12). The latter has an important effect on humoral response to one vaccine dose, as we have observed: in our study, 91% of dialysis patients (11/12) with prior SARS-CoV-2 infection developed detectable anti-RBD IgG.

Longitudinal follow-up of our cohort is ongoing, and additional patients are being recruited for this study as vaccinations roll out across the province. Whether humoral responses improve after a second dose and at what interval, and whether cellular immune responses develop after single or multiple doses in this population require further study. Tracking the rate symptomatic SARS-CoV-2 infection after vaccination in this and other studies will also aid greatly in our assessemnt of vaccine efficacy.

Our study has several strengths. We were able to obtain serial plasma samples before and after vaccination in all patients, and include both healthy individuals and dialysis patients with natural COVID-19 infection as controls.We were also reliably able to determine history of SARS-CoV-2 infection based on both pre-vaccination anti-RBD IgG and serial PCR data from weekly surveillance testing. (Pre-vaccination anti-RBD IgG and anti-N has sensitivity of 80% for prior SARS-CoV-2 detected by historical PCR). We recognize the limations of this data. We did measure T cell responses, follow-up is as yet short, and the small sample size does not allow us to identify patient characteristics predictive of humoral response or determine clinical efficacy.

While it is reassuring that the BNT162b2 vaccine is well-tolerated in dialysis patients, its efficacy remains uncertain. A single dose failed to elicit a humoral immune response in the majority of SARS-CoV-2 naïve patients even after prolonged observation, and in those with prior SARS-CoV-2 infection, the response was delayed. Whether HD patients develop effective T cell responses requires further study. Until then, we advise the second dose of the BNT162b2 be administered to all HD patients at the recommeded 3-week time interval, and that rigorous SARS-CoV-2 infection prevention and control measures be continued in dialysis units until vaccine efficacy is proven(13).

## Data Availability

De-identified aggregate data is available upon request to the lead author, RS, provided the necessary institutional approvals are received. Further information on the assays used and requests for reagents should be directed to and will be fulfilled by AF.

## Acknowledgements

This work is submitted by the authors on behalf of the COVID-19 Study Team of the *Réseau Rénal Québécois*. The authors thank the participants for donating their plasma samples. We are grateful to our research coordinators, Ms. Norka Rios, Ms. Marie-Line Caron, and Ms. Guylaine Marcotte. We thank Dr. Nick Bertos and the Research Institute of the McGill University Health Center for sample processing and storage. RS, RG, ACNF, and WBS are supported by the Fonds de Recherche du Québec – Santé (FRQS) Clinician-Researcher Awards. AF is the recipient of Canada Research Chair on Retroviral Entry no. RCHS0235950-232424. This study was funded by a Canadian Institutes of Health Research (CIHR) Rapid Research COVID-19 funding opportunity. This work was also supported by le Ministère de l’Économie et de l’Innovation du Québec, Programme de soutien aux organismes de recherche et d’innovation to AF, by the Fondation du CHUM and by a CIHR Foundation Grant #352417 to AF.

## Disclosures

None.

## Data and Materials

De-identified data is available upon request to the lead author, Rita Suri. Further information and requests for reagents should be directed to and will be fulfilled by AF.

## Notes

**Funding:** This study was funded by a Canadian Institutes of Health Research (CIHR) Rapid Research COVID-19 funding opportunity. This work was also supported by le Ministère de l’Économie et de l’Innovation du Québec, Programme de soutien aux organismes de recherche et d’innovation to AF, by the Fondation du CHUM and by a CIHR Foundation Grant #352417 to AF.

### Competing Interest Statement

The authors have declared no competing interest.

### Funding Statement

This study was funded by the Canadian Institutes of Health Research (CIHR) Rapid Research COVID-19 funding opportunity. This work was also supported by le Ministère de l'Économie et de l'Innovation du Québec, Programme de soutien aux organismes de recherche et d'Innovation to AF, by the Fondation du CHUM and by a CIHR Foundation Grant #352417 to AF. RS, RG, ACNF, and WBS are supported by the Fonds de Recherche du Québec Santé (FRQS) Clinician-Researcher Awards. AF is the recipient of Canada Research Chair on Retroviral Entry no. RCHS0235950-232424.

### Author Declarations

Comité d'éthique de la recherche clinique du CR-CHUM and l'Hôpital Sacre-Coeur

### Summary of Updates

This manuscript has been updated with 8-week data. Figure 1 and Table 1 have been revised.

